# GLP-1 Receptor Agonists and Cardiovascular Events During Androgen Receptor Pathway Inhibitor Therapy

**DOI:** 10.64898/2026.05.26.26353962

**Authors:** Katelyn M Atkins, Nikhil Chakravarty, Maria Oorloff, Giana Grigsby, Irfan Khan, Mitchell Kamrava, Andriana Nikolova, Anja Karlstaedt, Cody Ramin, Leslie K. Ballas

## Abstract

**Background:** Androgen receptor pathway inhibitors (ARPIs) have transformed the treatment of high risk and metastatic prostate cancer, though are associated with increased cardiovascular risk. Glucagon-like peptide-1 receptor agonists (GLP-1 RAs) have been shown to reduce cardiovascular events in non-cancer populations, but their role in patients receiving ARPIs is unclear.

**Methods:** Retrospective analysis of 120 men with PC treated with ARPIs between 2015-2025 with any GLP-1 RA exposure. The time of GLP-1 RA use was categorized relative to ARPI initiation (pre- vs post-ARPI). Cumulative incidences for major adverse cardiac events (MACE) any grade 2 or greater cardiac common terminology criteria for adverse events (CTCAE) were estimated. Fine-Gray regressions were performed (non-cardiac death as a competing risk).

**Results:** The median follow-up was 2.3 years (interquartile range [IQR] 1.3-3.7). The median age was 72 years (IQR 66-78). Atherosclerotic cardiovascular disease (ASCVD) was present in 45.0% (n=54). Overall, 55.0% (n=66) initiated GLP-1 RA therapy prior to ARPI and 45.0% (n=54) after ARPI initiation, with a median duration of GLP-1 RA use of 4.0 years (IQR, 2.3-7.0) and 1.3 years (IQR, 0.6-2.1), respectively. Four patients experienced MACE, including three coronary revascularizations and one ischemic stroke. 25 patients experienced at least one grade 2 or greater cardiac event, most commonly arrhythmia (n=20) and thromboembolic disease (n=11). The 2-year cumulative incidence of MACE and grade ≥2 cardiac events was 1.7% and 16.1%, respectively. Adjusting for pre-existing cardiovascular risk, GLP-1 RA duration, and pre- ARPI androgen deprivation therapy use, GLP-1RA use prior to ARPI initiation (vs. after ARPI start) was associated with reduced risk of grade ≥2 cardiac events (subdistribution hazard ratio 0.26, 95% CI 0.08-0.91; p=0.036).

**Conclusion:** GLP-1 RA use prior to ARPI initiation was associated with reduced risk of cardiac events, suggesting that earlier metabolic optimization may influence cardiovascular outcomes. These hypothesis-generating findings support investigation of early GLP-1 RA initiation as a potential cardiovascular risk mitigation strategy during ARPI therapy.

## INTRODUCTION

Cardiovascular disease is a major cause of morbidity and mortality among men with prostate cancer (PC)^1^. Androgen receptor pathway inhibitors (ARPIs) have transformed the treatment of high-risk patients with M0-M1 PC^2^, though are associated with adverse cardiometabolic effects, including increased central adiposity, elevated low-density lipoprotein levels, impaired glycemic control, and major adverse cardiac events (MACE)^3^. Glucagon-like peptide-1 receptor agonists (GLP-1 RAs) stimulate insulin secretion, reduce adiposity, modulate inflammation, and have been shown to reduce cardiovascular events in non-cancer populations^4^, but their role in patients receiving ARPIs is unclear. We evaluated whether GLP-1 RA use and timing relative to ARPI initiation were associated with cardiovascular events.

## METHODS

Retrospective analysis of 120 men with PC treated with ARPIs between 2015-2025 with any GLP-1 RA exposure. The time of GLP-1 RA use was categorized relative to ARPI initiation (pre-vs post-ARPI). Composite cardiovascular events after ARPI initiation were abstracted manually and adjudicated, including major adverse cardiac events (MACE: myocardial infarction, unstable angina, heart failure, coronary revascularization, ischemic stroke, and cardiovascular death) and any grade ≥2 cardiac common terminology criteria for adverse events (CTCAE). Cumulative incidences were estimated, and Fine-Gray regressions were performed (non-cardiac death as a competing risk). This study was conducted under approval from the Cedars-Sinai Medical Center IRB (CSMC STUDY00004097) and adhered to STROBE reporting guidelines.

## RESULTS

The median follow-up was 2.3 years (interquartile range [IQR] 1.3–3.7). The median age was 72 years (IQR 66–78); 63.3% (n=76) had metastatic disease, 61.4% (n=72) were on androgen deprivation therapy (ADT) prior to ARPI therapy. Cardiovascular comorbidities were common, including hypertension (84.2%, n=101), hyperlipidemia (81.2%, n=98), diabetes (70.0%, n=84), and atherosclerotic cardiovascular disease (ASCVD; 45.0%, n=54), though only 59.3% (32/54) of patients with ASCVD were on standard-of-care statin therapy at the time of ARPI initiation. Overall, 55.0% (n=66) initiated GLP-1 RA therapy prior to ARPI (median interval of 1.6 years [IQR, 0.6-5.2 years]) and 45.0% (n=54) after ARPI initiation (median interval 1.2 years [IQR, 0.4-2.8 years]), with a median duration of GLP-1 RA use of 4.0 years (IQR, 2.3-7.0) and 1.3 years (IQR, 0.6-2.1), respectively. Patients with GLP-1 RA exposure prior to ARPI were more likely to have hypertension, diabetes, heart failure, and be on statin therapy (p<0.05) (Table).

Four patients experienced MACE, including three coronary revascularizations and one ischemic stroke. 25 patients experienced at least one grade ≥2 cardiac event, including arrhythmia (n=20), thromboembolic disease (n=11), valvular heart disease (n=10), coronary events (n=9), and heart failure (n=6). The 2-year cumulative incidence of MACE and grade ≥2 cardiac events was 1.7% (95% confidence interval [CI] 0.3-5.4%) and 16.1% (95% CI 9.6-24.1%), respectively. The median interval from start of ARPI to first MACE or grade ≥2 CE was 2.0 years (IQR, 0.1-5.8 years) and 1.3 years (IQR, 0.8-2.5 years), respectively. Adjusting for age, hyperlipidemia, pre-existing ASCVD, statin use, GLP-1 RA duration, and pre-ARPI ADT use, GLP-1RA use prior to ARPI initiation (vs. after ARPI start) was associated with reduced risk of grade ≥2 cardiac events (subdistribution hazard ratio 0.26, 95% CI 0.08-0.91; p=0.036). The corresponding 2-year grade ≥2 cardiac event rates were 11.3% (95% CI 4.0-22.7%) with pre-ARPI GLP-1 RA use vs 21.5% (95% CI 11.5-33.5%) with post-ARPI initiation GLP-1 RA use, respectively (Figure).

**Figure 1.**
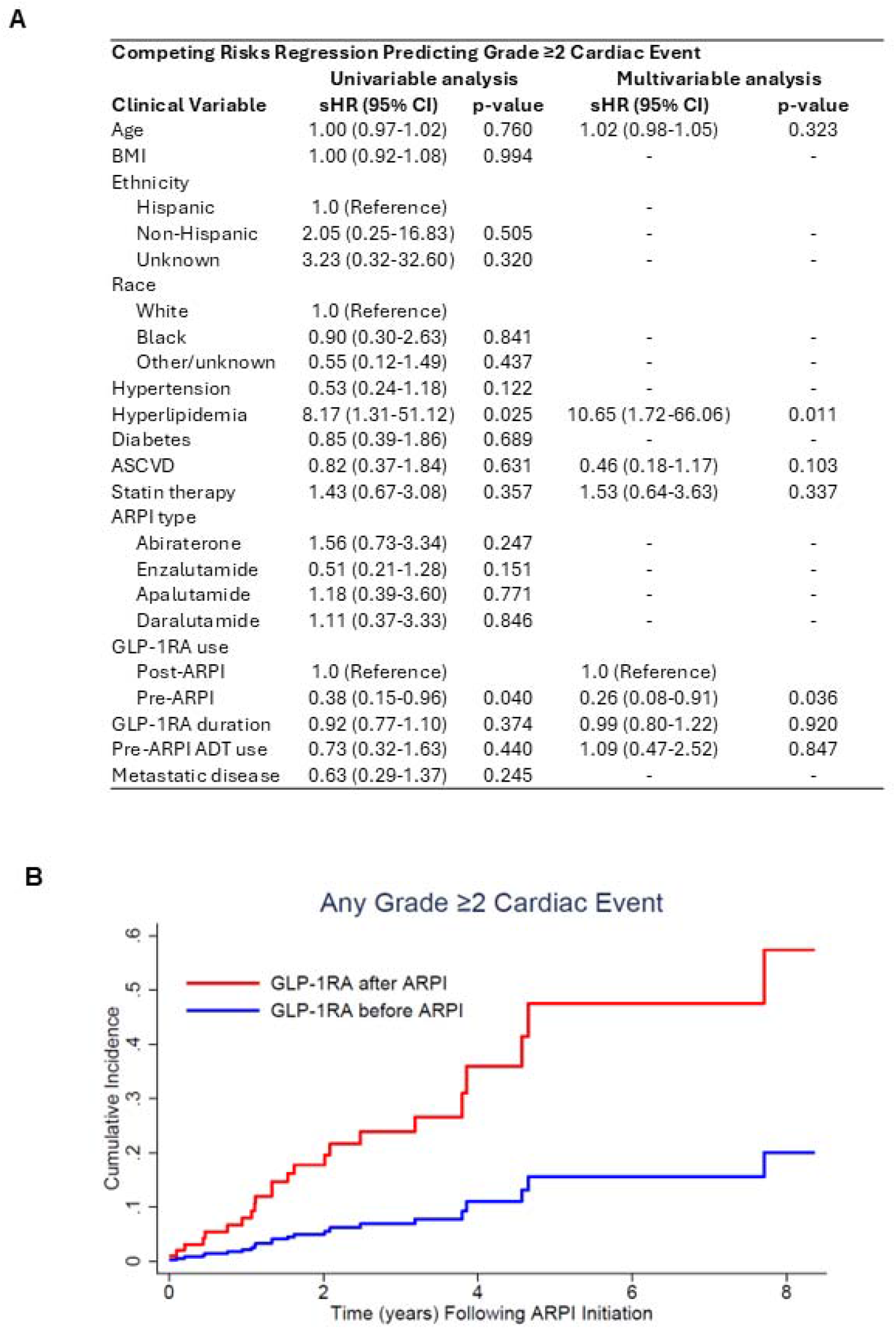
(A) Competing risks regression analysis predicting any grade ≥2 cardiac events. (B) Cumulative incidence estimates of any grade ≥2 cardiac events, stratified by GLP-1 receptor agonist (GLP-1 RA) use before or after androgen receptor pathway inhibitor (ARPI) therapy initiation. Abbreviations: sHR, subdistribution hazard ratio; BMI, body mass index; ASCVD, atherosclerotic cardiovascular disease.

## DISCUSSION

We observed that men with PC receiving ARPI therapy harbored substantial baseline cardiovascular risk, with notable statin underutilization despite high ASCVD prevalence, consistent with existing clinical trial data^5^. GLP-1 RA use prior to ARPI initiation was associated with reduced risk of cardiac events, suggesting that earlier metabolic optimization may influence cardiovascular outcomes. Together, these hypothesis-generating findings support investigation of early GLP-1 RA initiation as a potential cardiovascular risk mitigation strategy during ARPI therapy.

GLP-1 RAs show significant promise for reducing cardiovascular risk burden in patients with cancer. Early studies using the TriNetX Global Database demonstrate reduced risk of MACE and all-cause mortality in patients with cancer and diabetes^6^, including those treated with immune checkpoint inhibitors^7^, with monoclonal gammopathy of undetermined significance and diabetes^8^, and breast cancer and diabetes^9^, though data in PC is scarce. Further, given the swift adoption of GLP-1 RAs in populations with pre-existing cancer and without diabetes^10^, there is an urgent need for prospective studies to investigate the potential impact of GLP-1 RAs on cardiometabolic health and MACE outcomes. While no trials have yet reported such outcomes, active trials are evaluating GLP-1 RAs in PC patients receiving ADT, including IMPACT-ADT (NCT07202247), a randomized study focused on cardiometabolic health, while other studies primarily assess metabolic risk-factor modification rather than cardiovascular events (NCT06908694).

This study is limited by its small sample size, limited number of MACE, its retrospective design, and potential bias from unmeasured confounding variables. Nonetheless, these findings are suggestive that early GLP-1 RA use may augment existing cardiovascular risk reduction strategies in PC patients treated with ARPIs and are worthy of further investigation.

## Data Availability

The clinical data that support the findings of this study are available upon reasonable request by a qualified investigator under a data use agreement and with appropriate ethical oversight. The clinical data are not publicly available due to institutional review board restrictions regarding human subject research.

---

**Table 1.**
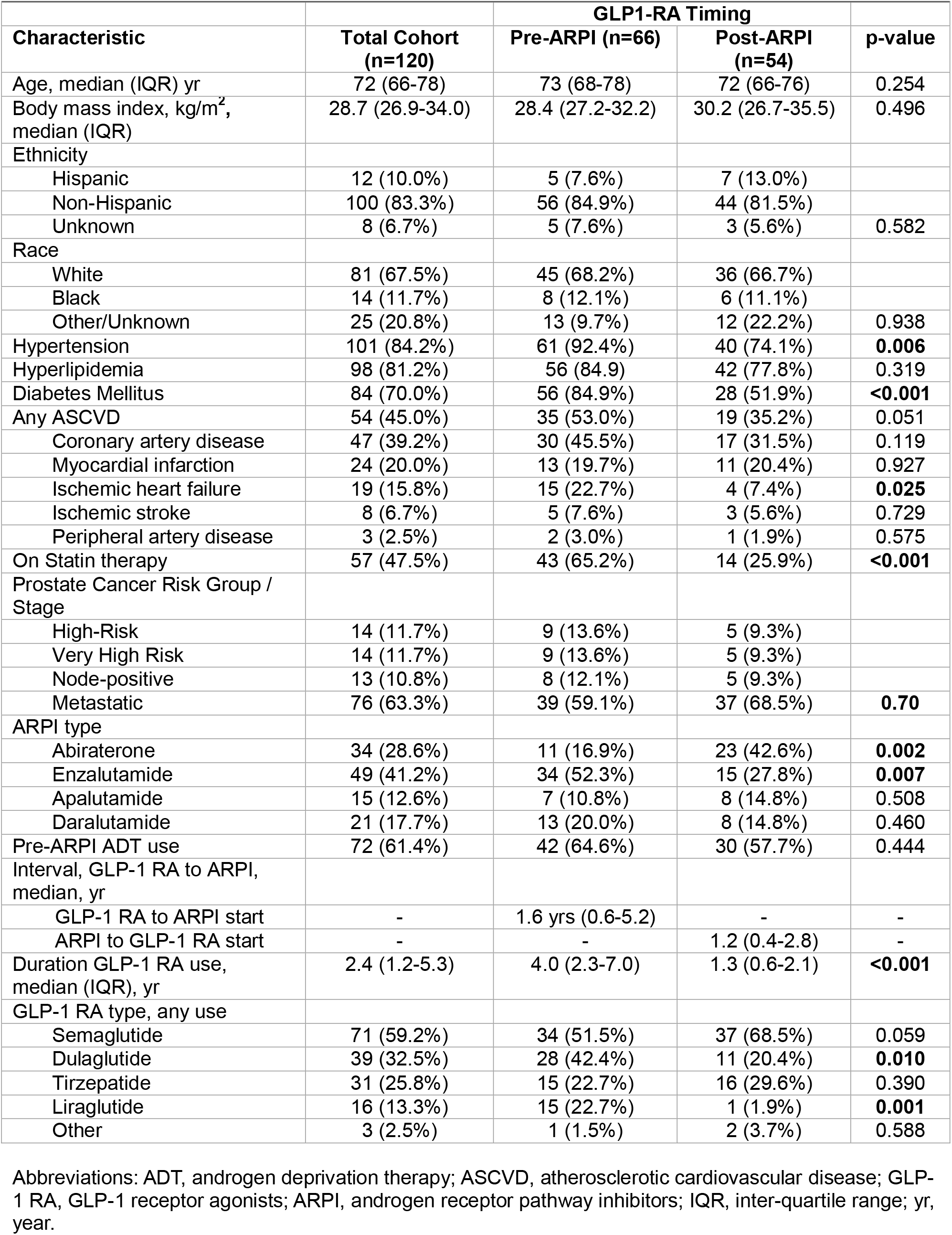
Baseline Clinical Characteristics.

